# Investigating the Relationship Between Radiotherapy Dose on the neurogenic niches and Overall Survival in NSCLC Brain Metastases

**DOI:** 10.1101/2023.05.10.23289385

**Authors:** Fia Cialdella, Danique E. Bruil, A.T.J. van der Boog, Steven H.J Nagtegaal, F.Y.F. de Vos, J.J.C. Verhoeff, Szabolcs David

## Abstract

**Background:** Non-small cell lung cancer (NSCLC), the most common type of lung cancer, often leads to brain metastases (BMs) with a poor prognosis. Radiotherapy is the main treatment for BMs, which despite decades of development, still results in radiation of healthy tissue. Neural stem cells (NSCs), crucial for the establishment and preservation of the nervous system, are sensitive to radiation, therefore radiation damage to NSCs may affect overall survival (OS). NSCs are primarily located in the subventricular zone (SVZ) and the subgranular zone (SGZ) within the hippocampus (HPC). Our study aims to evaluate the impact of radiotherapy dose on NSCs on OS in patients with BMs from NSCLC.

**Methods:** We have retrospectively included 138 NSCLC patients with BMs, irradiated at a single academic institute. NSC regions were delineated on the non-enhanced T1 MR images with CAT12 and SPM. The association between regional mean doses in the SVZ and HPC and OS was examined using a Cox regression model. Additionally, survival differences between lesion contact and no direct contact with SVZ and HPC were investigated with Kaplan-Meier (KM) analysis.

**Findings:** Multivariable Cox regression of dose on the SVZ and HPC showed a significant negative correlation, with a hazard ratio (HR) of 1.366 (p = 0.041 [95% (CI) 1.013– 1.842]) and 1.194 (p = 0.037 [95% CI 1.010 – 1.411]), respectively. KM analysis did not find a relationship between lesion contact with NSC-regions and OS.

**Interpretation:** Radiotherapy dose on the neurogenic niches is correlated with poorer OS and we found no association between direct lesion contact to NSC-regions and OS. We recommend further investigation into the impact of radiation on OS and neurocognitive function in a prospective study design in order to develop treatment approaches that minimize the potential harm to NSC’s while maximizing effectiveness.

**Funding:** Received no funds, grants, or support.

## BACKGROUND

### Lung cancer and brain metastases

The majority of patients with lung cancer are diagnosed with Non-Small Cell Lung Cancer (NSCLC), of which Lung Adenocarcinoma (LUAD ∼50%) and Lung Squamous Cell Carcinoma (LUSC ∼40%) are the most common types.^1^ A frequent complication of NSCLC is the development of brain metastases (BMs). The incidence has increased recent years due to more sensitive detection methods for BMs, longer life expectancy of cancer patients due to improved systemic therapy against the primary tumor, and because of an ageing population being more prone to developing cancers.^2^ The exact prevalence of BMs in NSCLC is unknown, with studies reporting a risk rate of 10% to 36% to develop BMs during the disease course.^3–6^ Overall survival (OS) of these patients varies from median 2 to 9 months depending on type of treatment.^7,8^ BMs are treated by either local radiotherapy (RT) or a combined treatment of surgical resection and subsequent RT. The aim of local therapy is to improve OS and to alleviate symptomatic progression of the BMs.^9^ The preferred type of radiotherapy is stereotactic radiosurgery (SRS), in which targeted radiation is delivered with high precision to the BMs - usually in a large, single fraction - to improve local tumor control. The prescribed SRS dose is calculated by the volume or axial size of the BMs based-on contrast enhanced T1-weighted MR imaging, while keeping the presumed risk of radiation damage to the surrounding tissue in mind.^10^ Whole-brain radiotherapy (WBRT) is also used to treat multiple BMs (e.g. more than 10), as prophylactic treatment of microscopic BMs in Small Cell Lung Cancer or as symptomatic treatment in terminal stage of the disease.^11^ However, due to this risk of toxicity, SRS is preferred over WBRT.^12^

Previous studies on general cranial radiotherapy (RT) indicate that RT can damage healthy brain tissue, leading to atrophy or damage that may subsequently result in cognitive impairment or other neurological disorders.^13,14^

Radiation therapy that irradiates neural stem cells (NSCs) can potentially have a negative impact on both OS and Quality of Life (QoL) due to the presumed effects on the repair of radiation-induced damage to the brain.^15^

### Neurogenic niches in adults

NSCs are located in the so called *neurogenic niches*, which involve two separate regions in the adult brain: (1) the subventricular zone (SVZ), situated underneath the ependymal layer of cells that line the ventricles and (2) the subgranular zone (SGZ), part of the hippocampal dentate gyrus.^16^ NSCs induce neurogenesis, which is the process of generating new neurons from NSCs. This process allows cell replacement after neuron loss triggered by brain injury (e.g. due to tumor growth, surgery or radiotherapy).^17^ Additionally, adult NSCs, particularly in the hippocampus (HPC) also play a role in the regulation of several cognitive proccesses.^18^ Irradiation of the neurogenic niches is hypothesized to play a role in reducing adult neurogenesis in two ways: inducing acute apoptosis in dividing cells and reducing the production of new neurons by decreasing the pool of mitotic NSCs.^19^

A recent study by our group examined the relationship between tumor contact and irradiation of the neurogenic niches^20^ in glioma patients, where we showed that a tumor which invades the SVZ region has decreased survival compared to patients without SVZ-tumor invasion. No association was observed for hippocampus-tumor invasion. We also found that irradiation of the neurogenic areas leads to a worser OS in patients with high grade glioma. According to the findings of this study, avoiding irradiation of the neurogenic niches in patients with glioma may result in a better OS.

The survival of lung cancer patients is increasing due to improved therapies such as molecularly targeted therapies ^5^ and the incidence of BMs is rising.^2^ To extend the survival of this patient group through appropriate radiotherapy and minimize the negative impact on their QoL, further research is necessary. Therefore, this retrospective study aims to investigate the effect of radiotherapy dose on neurogenic niches on OS in a cohort of NSCLC patients with BMs. Our hypothesis is that any dose on the neurogenic niches or overlap between lesions and neurogenic niches would result in lower OS.

## MATERIAL AND METHODS

### Patient Selection and clinical data collection

We retrospectively selected patients with BMs from NSCLC, who underwent SRS at a single academic center, the UMC Utrecht (UMCU), between December 2014 and April 2020. All patients have been referred from the UMC Utrecht and surrounding general hospitals in the Utrecht region, where they were treated for NSCLC. Inclusion criteria were: patients diagnosed with NSCLC, age ≥18 years and accessible RT planning and dosimetry data. Patients with a karnofsky performance status (KPS) of less than 70 were only included if there was an expectation of neurological improvement as an effect of stereotactic radiosurgery. Exclusion criteria were neuro endocrine pathology and prior intracranial radiotherapy. OS was monitored until June 28^th^ 2022 and censored afterwards. Clinical data was extracted from the electronic health record (HiX, Chipsoft, The Netherlands) or from referral data and included demographics (age, gender) as well as clinical information such as type of NSCLC, number and location of BMs, and prior systemic therapies (i.e., chemotherapy, immunotherapy and targeted therapy). Type of lung cancer was divided in (1) LUAD, including undifferentiated large cell carcinoma, and (2) LUSC. The WHO performance status was calculated based on the KPS by means of the oncology practice tool.^21^ Gross total volume (GTV) and planning target volume (PTV) were delineated for clinical purposes by a radiation oncologist. The UMC Utrecht Medical Ethics Research Committee assessed the study protocol and waived the need for informed consent (METC 18-273).

### Image processing and segmentation

Briefly, patients underwent pre-treatment MR scans on a 3T Philips scanner (Philips Healthcare, Best, The Netherlands) as part of standard clinical care. We collected the 3D turbo-spin echo (TSE) T1-weighted MR images without contrast enhancement, with the following parameters: TR = 8.1 ms, TE = 3.7 ms, flip angle = 8°, voxel resolution 1 × 0.96 × 0.96mm. The planning CT scans were acquired on a Brilliance Big bore scanner (Philips Medical Systems, Best, The Netherlands), with a tube potential of 120 kVp, using a matrix size of 512 × 512 and 0.65 × 0.65 × 3.0 mm voxel size. While automated segmentation of MR images with common neuroimaging software, e.g.: FSL, SPM and FreeSurfer has great performance, it is often challenging and lead to unsatisfactory results when mayor abnormalities are present in the brain, such as BM lesions. In order to facilitate the segmentation, first we replaced all the abnormal tissues, that is edema, tumor cavity, etc., with healthy appearing, but fake tissue, which process is also called virtual brain grafting (VBG).^22^ This allowed the automatic delineation of the SVZ and hippocampus regions on the VBG-enhanced T1 MR images with CAT12. The readers are kindly referred to our previous work at *Bruil et al (2022)*.^20^, which describe the fine details of all image processing. An example of delineation and radiotherapy plan is presented in **Figure 1**. After the process finished, regions and labels covering the fake tissue were ignored from further analysis, therefore only the non-affected parts of SVZ and hippocampus were considered. Mean dose per SVZ and hippocampi labels were calculated for further analyses, while SVZ/hippocampi to lesion contact was defined when SVZ/hippocampi labels from the aforementioned segmentation process were part of the replaced fake tissue. In case SVZ/hippocampi labels were outside of the fake tissue, it is considered no lesion contact.

**Figure. 1:**
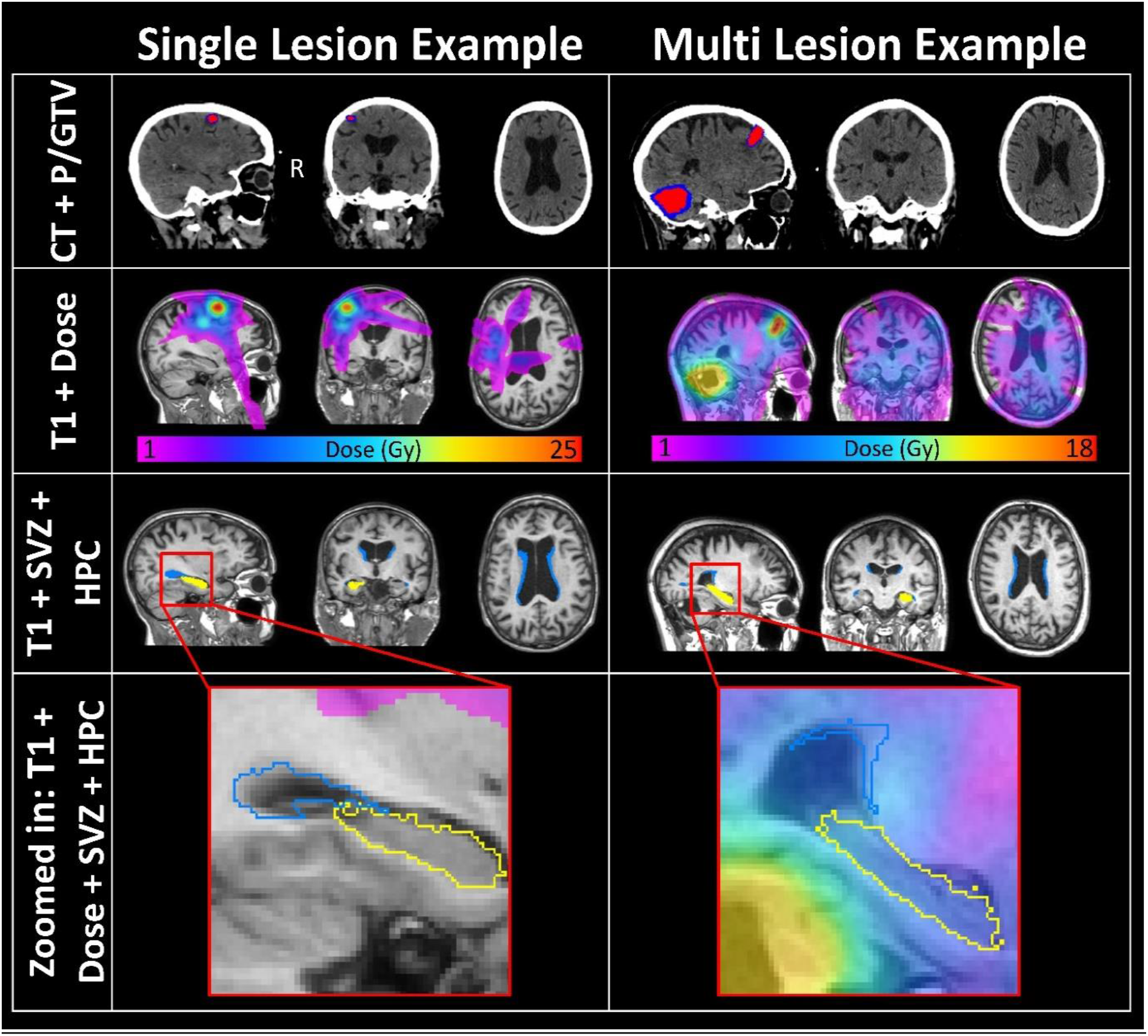
An example of delineation and radiotherapy. Two example patients’ radiotherapy plan and the SVZ, hippocampus delineations, respectively. The patient on the left has a single brain metastatic lesion, which is distal from the both SVZ and hippocampus, while the patient of the right has two BM lesions of which one is proximal the neurogenic niches. The top two rows show the CT scan along with the PTV and CTV in blue and red respectively as well as the dose distribution. The bottom two rows show the SVZ and a hippocampus segment. Radiological view: left is right.

### Dose calculation

Typically, a single fraction of RT was prescribed. In case of multiple fractions given instead of a single treatment, the dose was recalculated in single fraction equivalent dose (SFED). We defined SFED as the dose delivered in a single fraction that would have the same biologic effect as the dose-fractionation scheme, according to the method by *Park et al*.*(2008)*^23^

### Statistical Analyses

The relationship between OS, lesion contact and mean dose per neurogenic niche region were analyzed with IBM SPSS (for Windows, version 25). The primary outcome of this study was OS, defined as months after the RT until date of death. In this study, we utilized Kaplan-Meier (KM) analyses to investigate the impact of lesion contact with the neurogenic niches on OS in patients. Specifically, we compared patients who had lesion contact with the neurogenic niches to those who did not. Additionally, we conducted multivariable Cox-regression analyses, corrected for the covariates outlined in the results section, to analyze OS. We set statistical significance at p <0.05. Furthermore, we used the Variance Inflation Factor (VIF) to evaluate the presence of multi-collinearity among all covariates for each independent variable.

## RESULTS

### Participants

From December 2014 to April 2020, we identified 802 patients who underwent RT for BMs at the UMCU. Among these patients, 389 had previously received intracranial RT, such as SRS, WBRT or prophylactic cranial irradiation (PCI), and were therefore excluded from the study. Of the 413 patients who met the inclusion criteria, 141 were diagnosed with BMs from non-small cell lung cancer (NSCLC). After excluding 3 patients with neuroendocrine pathology, the final study cohort comprised 138 patients. The patient inclusion flowchart is shown in **Figure 2**. Baseline patient and treatment characteristics are shown in **Table 1**. The median age was 67 years (interquartile range 60 -74) and the male-female distribution was even. 28 (20%) patients underwent surgery prior to RT of whom 21 patients had a complete macroscopic resection of one lesion, next to other unresected BMs. A total of 29.7% of the patients received a concurrent combination of chemotherapy and other systemic treatments, 28.4% was treated by chemotherapy alone and 19.1% received immunotherapy/target therapy only.

**Table 1:**
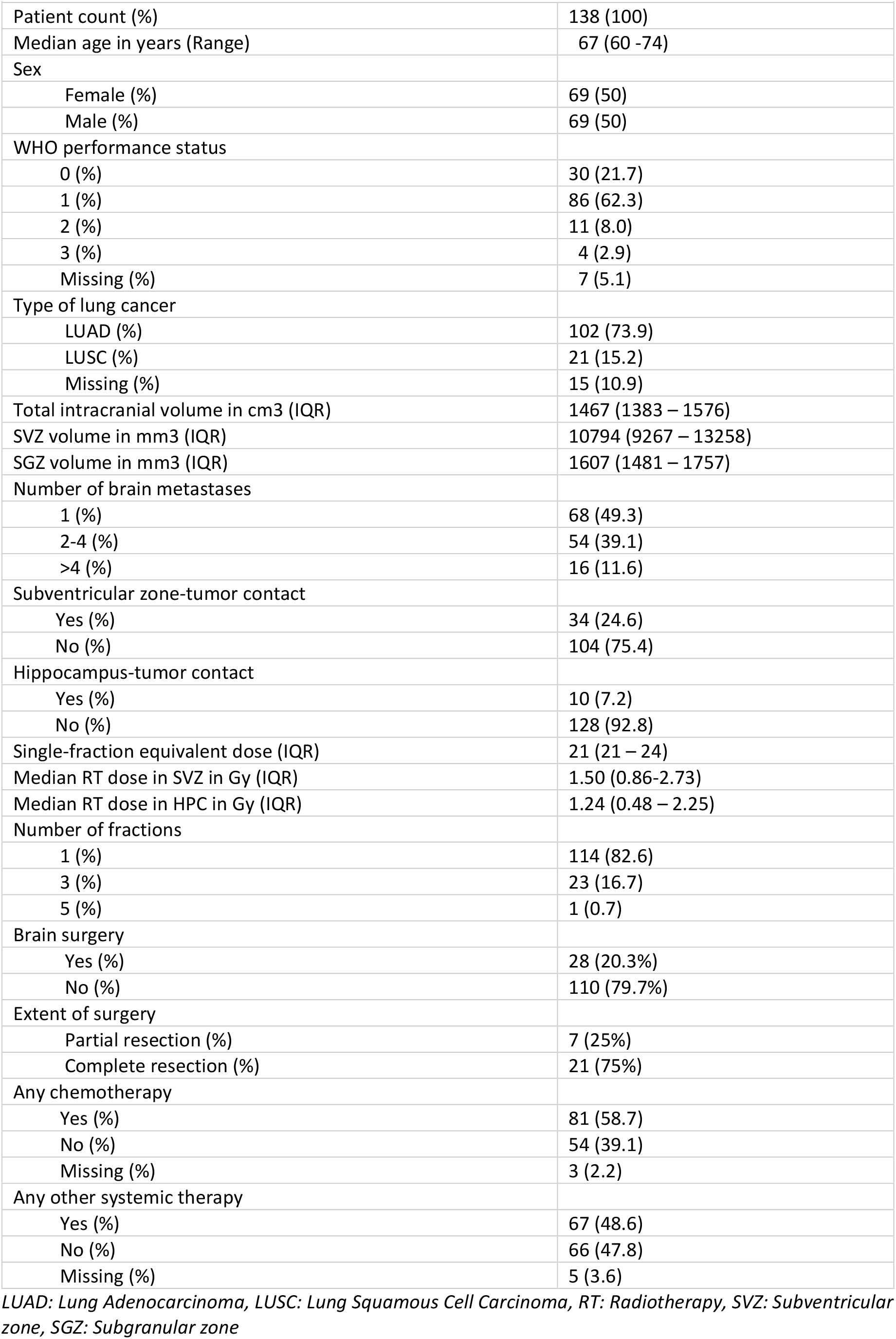
Baseline patient and treatment characteristics.

| **Table 1: Baseline patient and treatment characteristics** |  |
| --- | --- |
| Patient count (%) | 138 (100) |
| Median age in years (Range) | 67 (60 -74) |
| Sex |  |
| Female (%) | 69 (50) |
| Male (%) | 69 (50) |
| WHO performance status |  |
| 0 (%) | 30 (21.7) |
| 1 (%) | 86 (62.3) |
| 2 (%) | 11 (8.0) |
| 3 (%) | 4 (2.9) |
| Missing (%) | 7 (5.1) |
| Type of lung cancer |  |
| LUAD (%) | 102 (73.9) |
| LUSC (%) | 21 (15.2) |
| Missing (%) | 15 (10.9) |
| Total intracranial volume in cm3 (IQR) | 1467 (1383 – 1576) |
| SVZ volume in mm3 (IQR) | 10794 (9267 – 13258) |
| SGZ volume in mm3 (IQR) | 1607 (1481 – 1757) |
| Number of brain metastases |  |
| 1 (%) | 68 (49.3) |
| 2-4 (%) | 54 (39.1) |
| >4 (%) | 16 (11.6) |
| Subventricular zone-tumor contact |  |
| Yes (%) | 34 (24.6) |
| No (%) | 104 (75.4) |
| Hippocampus-tumor contact |  |
| Yes (%) | 10 (7.2) |
| No (%) | 128 (92.8) |
| Single-fraction equivalent dose (IQR) | 21 (21 – 24) |
| Median RT dose in SVZ in Gy (IQR) | 1.50 (0.86-2.73) |
| Median RT dose in HPC in Gy (IQR) | 1.24 (0.48 – 2.25) |
| Number of fractions |  |
| 1 (%) | 114 (82.6) |
| 3 (%) | 23 (16.7) |
| 5 (%) | 1 (0.7) |
| Brain surgery |  |
| Yes (%) | 28 (20.3%) |
| No (%) | 110 (79.7%) |
| Extent of surgery |  |
| Partial resection (%) | 7 (25%) |
| Complete resection (%) | 21 (75%) |
| Any chemotherapy |  |
| Yes (%) | 81 (58.7) |
| No (%) | 54 (39.1) |
| Missing (%) | 3 (2.2) |
| Any other systemic therapy |  |
| Yes (%) | 67 (48.6) |
| No (%) | 66 (47.8) |
| Missing (%) | 5 (3.6) |

LUAD: Lung Adenocarcinoma, LUSC: Lung Squamous Cell Carcinoma, RT: Radiotherapy, SVZ: Subventricular zone, SGZ: Subgranular zone

**Figure 2:**
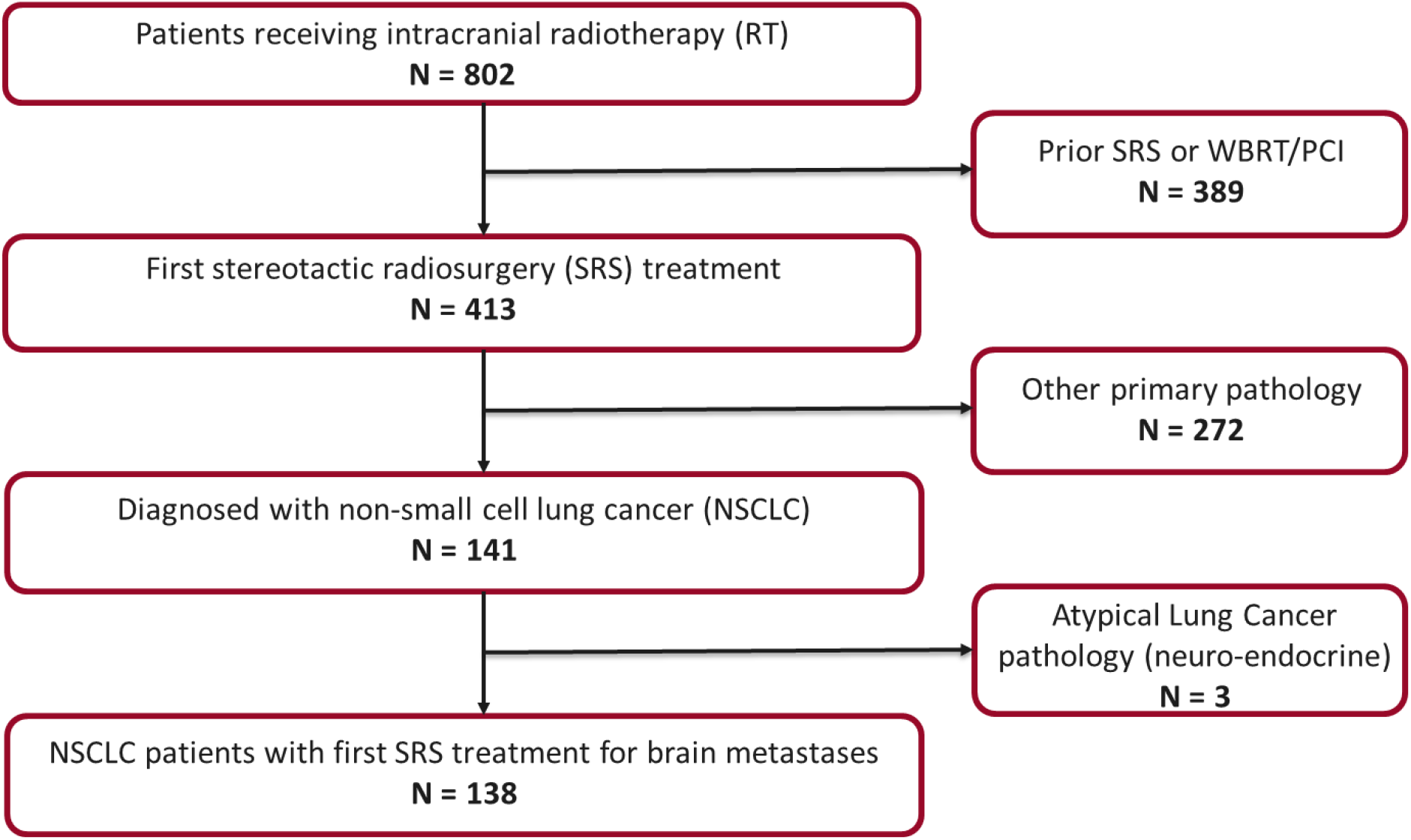
Flowchart of patient inclusion. RT: Radiotherapy, SRS: Stereotactic radiosurgery, WBRT: Whole brain radiotherapy, PCI: Prophylactic cranial irradiation, NSCLC; non-small cell lung cancer

### Survival Analysis

At time of censoring, 107 (78%) of the 138 patients within our cohort had deceased, with a median OS from start of RT of 8.4 months (range 0.23 – 64.7 months). In LUAD, we observed a median OS rate of 9.6 months and for LUSC 5.1 months, which were not significantly different (p = 0.461). As can be seen in **Figure 3** and **Table 2**, we found no association between OS and lesion contact with the SVZ or HPC in univariate, nor different multivariate setups.

**Table 2:** Univariable and multivariable Cox regression analysis of no tumor contact/tumor contact with the subventricular zone (SVZ) on overall survival (OS). Analyses with p < 0.05 are considered significant and bolded in the table.

|  |  |  |  |  |
| --- | --- | --- | --- | --- |
| Univariable: |  | p-value | 0.465 |  |
|  |  | HR | 1.177 |  |
|  |  | 95.0 % CI | Lower | 0.760 |
|  |  |  | Upper | 1.823 |
| Multivariable, corrected for: | Age, Sex, WHO performance status, type of lung cancer, SVZ volume | P-value | 0.336 |  |
|  |  | HR | 1.265 |  |
|  |  |  | 0.783 |  |
|  |  | 95.0% CI | Lower |  |
|  |  |  | Upper | 2.042 |
| Multivariable, corrected for: | Age, Sex, WHO performance status, type of lung cancer, SVZ volume, number of brain metastases, metastases elsewhere in the body, chemotherapy, any other systemic therapy, resection extent | p-value | **0.047** |  |
|  |  | HR | **1.670** |  |
|  |  | 95.0% CI | Lower | **1.006** |
|  |  |  | Upper | **2.773** |
| Multivariable, corrected for: | Age, Sex, WHO performance status, type of lung cancer, SVZ volume, number of brain metastases, metastases elsewhere in the body, chemotherapy, any other systemic therapy, resection extent, % PTV within the SVZ | p-value | 0.220 |  |
|  |  | HR | 1.472 |  |
|  |  | 95.0% CI | Lower | 0.794 |
|  |  |  | Upper | 2.727 |

**Figure 3:**
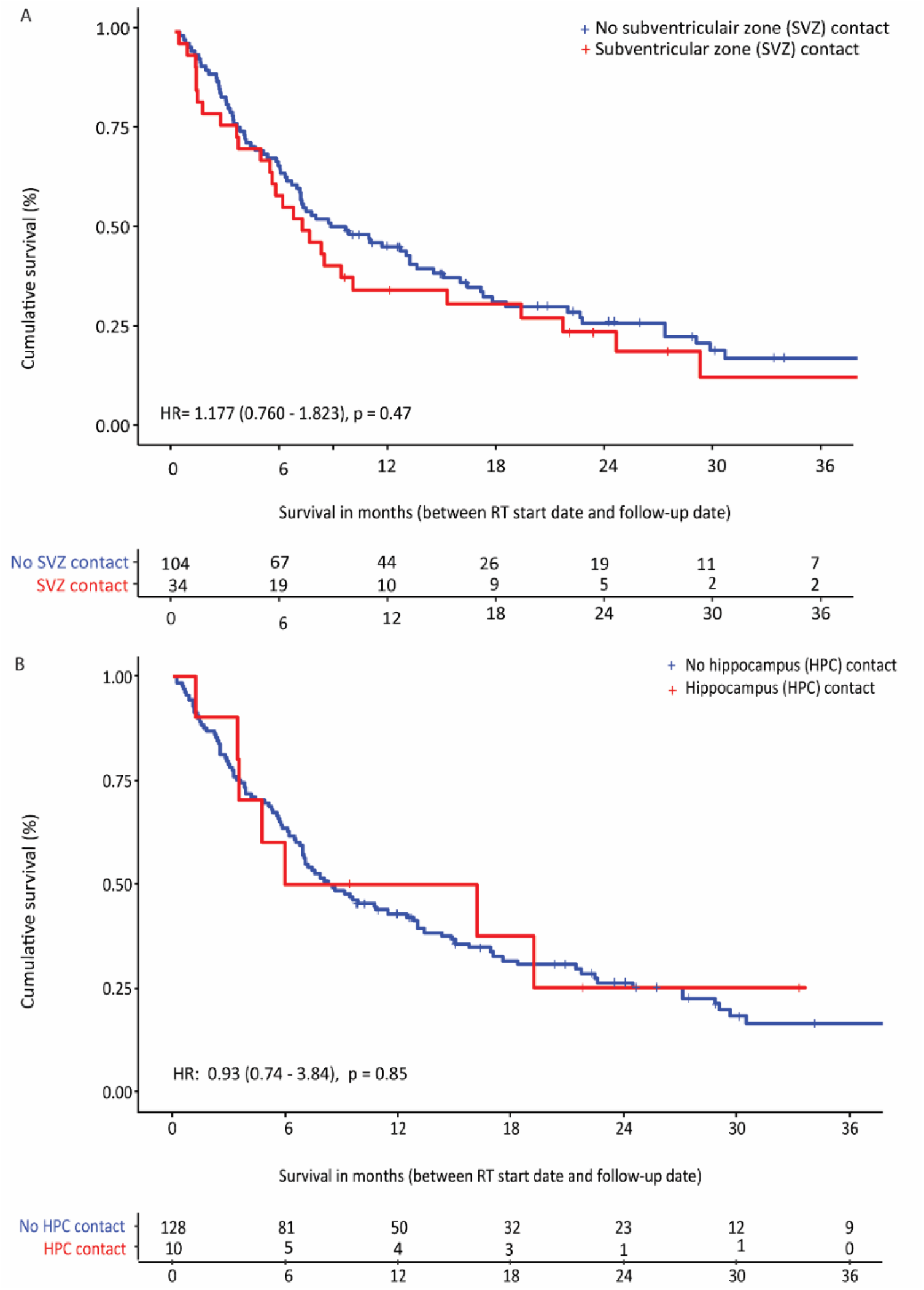
A) Kaplan-Meier overall survival according to tumor contact with the subventricular zone (SVZ) B) Kaplan-Meier overall survival according to tumor contact with the hippocampus (HPC). HR=hazard ratio. LUAD=lung adenocarcinoma. LUSC=lung squamous cell carcinoma

**Table 3:** Univariable and multivariable Cox regression analysis of no tumor contact/tumor contact with the hippocampus (HPC) on overall survival (OS). Analyses with p < 0.05 are considered significant and bolded in the table.

|  |  |  |  |  |
| --- | --- | --- | --- | --- |
| Univariable: |  | p-value | 0.852 |  |
|  |  | HR | 0.930 |  |
|  |  | 95.0% CI | Lower | 0.431 |
|  |  |  | Upper | 2.003 |
| Multivariable, corrected for: | Age, Sex, WHO performance status, type of lung cancer, SGZ volume | p-value | 0.209 |  |
|  |  | HR | 1.692 |  |
|  |  | 95.0 % CI | Lower | 0.745 |
|  |  |  | Upper | 3.843 |
| Multivariable, corrected for: | Age, Sex, WHO performance status, type of lung cancer, SGZ volume, number of brain metastases, metastases elsewhere in the body, chemotherapy, any other systemic therapy, resection extent | p-value | 0.157 |  |
|  |  | HR | 1.839 |  |
|  |  | 95.0% CI | Lower | 0.791 |
|  |  |  | Upper | 4.275 |
| Multivariable, corrected for: | Age, Sex, WHO performance status, type of lung cancer, SGZ volume, number of brain metastases, metastases elsewhere in the body, chemotherapy, any other systemic therapy, resection extent, % PTV within the SGZ | p-value | 0.163 |  |
|  |  | HR | 1.821 |  |
|  |  | 95.0% CI | Lower | 0.784 |
|  |  |  | Upper | 4.231 |

HPC: Hippocampus, SGZ: subgranular zone, PTV: Planned target volume.

**Table 4** and **5** show the results of the univariate- and multivariate Cox-regression between mean SVZ/HPC dose and OS, respectively. Both the radiotherapy dose on SVZ and HPC have a significant negative association with OS after adjustment for covariates with HR of 1.366 (p = 0.041 [95% confidence interval (CI) 1.013– 1.842]) and 1.194 (p = 0.037 [95% CI 1.010 – 1.411]), respectively.

**Table 4:**
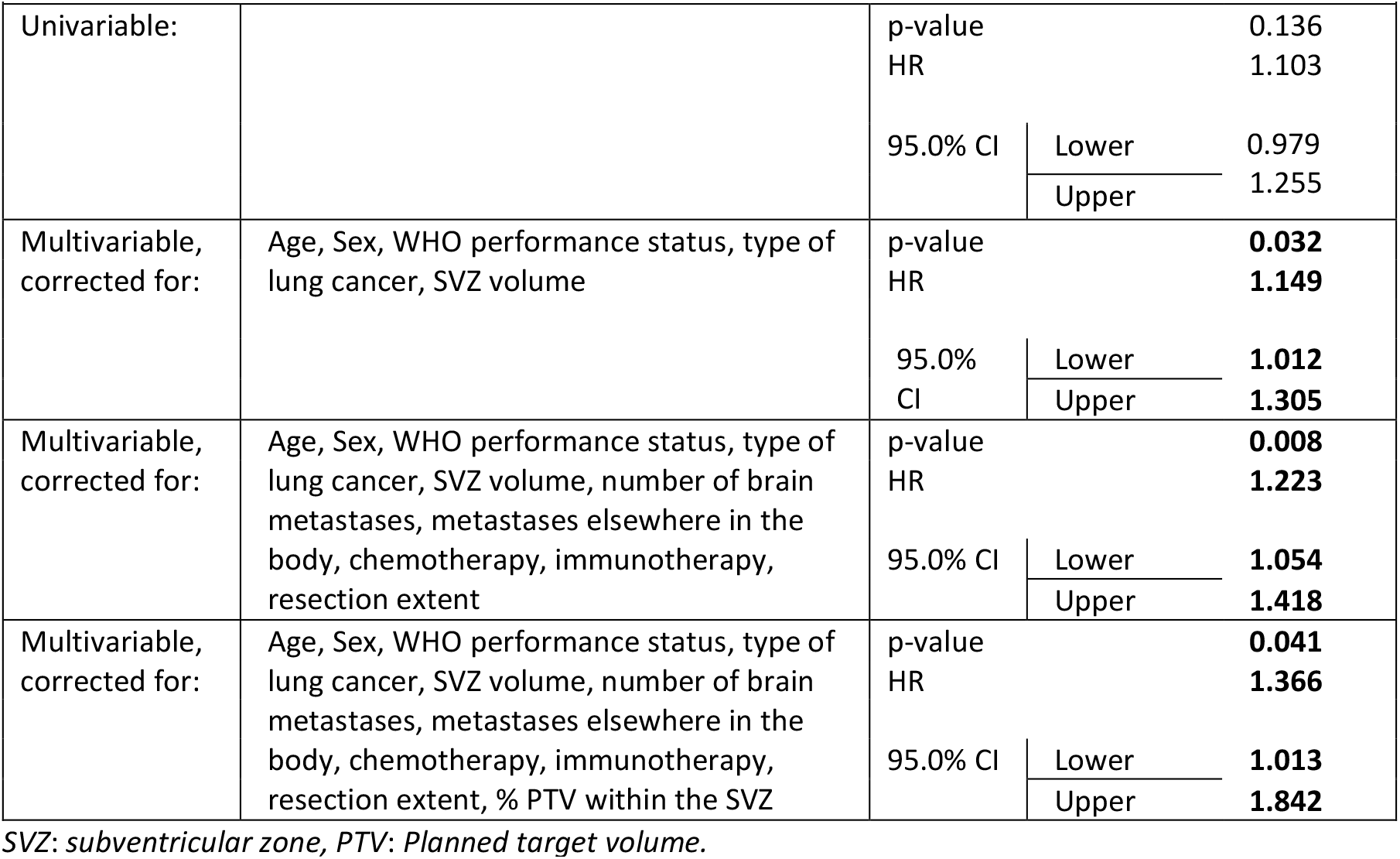
Univariable and multivariable Cox regression analysis of dose on the subventricular zone (SVZ) on overall survival (OS). Analyses with p < 0.05 are considered significant and bolded in the table.

|  |  |  |  |  |
| --- | --- | --- | --- | --- |
| Univariable: |  | p-value | 0.136 |  |
|  |  | HR | 1.103 |  |
|  |  | 95.0% CI | Lower | 0.979 |
|  |  |  | Upper | 1.255 |
| Multivariable, corrected for: | Age, Sex, WHO performance status, type of lung cancer, SVZ volume | p-value | **0.032** |  |
|  |  | HR | **1.149** |  |
|  |  | 95.0% CI | Lower | **1.012** |
|  |  |  | Upper | **1.305** |
| Multivariable, corrected for: | Age, Sex, WHO performance status, type of lung cancer, SVZ volume, number of brain metastases, metastases elsewhere in the body, chemotherapy, immunotherapy, resection extent | p-value | **0.008** |  |
|  |  | HR | **1.223** |  |
|  |  | 95.0% CI | Lower | **1.054** |
|  |  |  | Upper | **1.418** |
| Multivariable, corrected for: | Age, Sex, WHO performance status, type of lung cancer, SVZ volume, number of brain metastases, metastases elsewhere in the body, chemotherapy, immunotherapy, resection extent, % PTV within the SVZ | p-value | **0.041** |  |
|  |  | HR | **1.366** |  |
|  |  | 95.0% CI | Lower | **1.013** |
|  |  |  | Upper | **1.842** |

SVZ: subventricular zone, PTV: Planned target volume.

**Table 5:** Univariable and multivariable Cox regression analysis of dose on the hippocampus (HPC) on overall survival (OS). Analyses with p < 0.05 are considered significant and bolded in the table.

|  |  |  |  |  |
| --- | --- | --- | --- | --- |
| Univariable: |  | p-value | 0.087 |  |
|  |  | HR | 1.113 |  |
|  |  | 95.01% CI | Lower | 0.984 |
|  |  |  | Upper | 1.258 |
| Multivariable, corrected for: | Age, Sex, WHO performance status, type of lung cancer, SGZ volume | p-value | 0.058 |  |
|  |  | HR | 1.144 |  |
|  |  | 95.0% CI | Lower | 0.996 |
|  |  |  | Upper | 1.314 |
| Multivariable, corrected for: | Age, Sex, WHO performance status, type of lung cancer, SGZ volume, number of brain metastases, metastases elsewhere in the body, chemotherapy, any other systemic therapy, resection extent | p-value | 0.091 |  |
|  |  | HR | 1.146 |  |
|  |  | 95.0% CI | Lower | 0.978 |
|  |  |  | Upper | 1.341 |
| Multivariable, corrected for: | Age, Sex, WHO performance status, type of lung cancer, SGZ volume, number of brain metastases, metastases elsewhere in the body, chemotherapy, any other systemic therapy, resection extent, % PTV within the SGZ | p-value | **0.037** |  |
|  |  | HR | **1.194** |  |
|  |  | 95.0% CI | Lower | **1.010** |
|  |  |  | Upper | **1.411** |

HPC: Hippocampus, SGZ: subgranular zone, PTV: Planned target volume.

Our study showed that WHO performance status is correlated to OS.

## DISCUSSION

This retrospective study was designed to determine the effect of radiotherapy dose from the neurogenic niches on OS and if lesion contact with the SVZ or the HPC regions affect OS in patients with BMs from NSCLC.

Multivariate cox regression analysis showed a negative association between OS and radiotherapy dose on the SVZ or the HPC after adjusting for confounding variables. The SVZ plays a pivotal role in neurogenesis and brain repair mechanisms in adults, as reported by Capilla-Gonzalez et al. (2016).^15^ Therefore, it could be hypothesized that reduced brain repair capacity due to radiation-induced damage to the SVZ may contribute to the negative impact on OS. In addition, previous research has highlighted the role of adult NSCs of the HPC in regulating cognitive function such as learning and memory.^18^ Moreover, literature suggest that, independent of initial level of cognition, a faster decline of cognitive function is associated with a higher mortality. ^24^ Based on such functions, the importance of hippocampal avoidance to maintain neurocognitive function is established. In line with this, clinical trials already have supported this observation by demonstrating that hippocampal avoidance during whole-brain radiation therapy (HA-WBRT) can lead to a considerable decrease in cognitive decline.^25^ Currently, there is a limited number of investigations reporting the relationship between dose received by the SVZ regions in CNS tumors and neurocognitive decline after radiotherapy, which showed deterioration in cognition after higher radiation dose on the SVZ.^26^ Further research should be conducted to investigate this in more detail. Our ongoing ‘Cohort for patient-reported Outcomes, Imaging and trial inclusion in Metastatic BRAin disease’ (COIMBRA) trial *(ClinicalTrials*.*gov Identifier: NCT05267158)* is an example of such an initiative that can confirm the effect of radiation dose on the neurogenic niches on cognitive functioning of patients with BMs.

This current study showed no relationship between lesion contact with the neurogenic niches and OS of patients with BMs from NSCLC. Likely due to the sample size of the group with contact with the neurogenic niches, we were unable to determine whether the extent of contact has a significant impact. Previous research suggested that components of the neurogenic niches stimulate metastatic tumor growth ^27^, suggesting that tumor growth is inhibited when this component of the neurogenic niches is disrupted by radiation damage to the NSCs. Future research should aim at gaining more insight into the role of NSCs in metastatic pathways.

The current study adds significant value to the field of neuro-oncological imaging by providing novel insights into the impact of radiotherapy dose on the neurogenic niches in NSCLC patients with BMs. The study utilized high-quality imaging data and a specially developed atlas^20^ to enable accurate delineation of the regions of interest. Moreover, the patient population included in the study reflects the average patient with BMs from NSCLC, increasing the generalizability of the findings. These strengths enhance the potential clinical significance and impact of the study, providing a strong foundation for future research in this area.

There are several limitations to our study. Ideally, we had a more even distribution between the group with the patients with lesion contact with the SVZ or HPC and the patients without, as this would give our statistical analysis more power and would give us the opportunity to also investigate the subject in subgroups such as type of lung cancer and number of BMs. Nevertheless, previous studies already suggested that the overall risk for the incidence of metastases in the neurogenic niches is quite low. ^17,28^ It seems that our cohort is an accurate reflection of the population and lesion contact with the neurogenic niches is indeed less common. While the inclusion of patients referred from multiple hospitals allowed for a more diverse patient population in the study, one disadvantage is the occurrence of missing data regarding specific gene mutations and progression free survival (PFS).

Our study shows a median OS of 8.3 months in selected patients with BMs from NSCLC. Furthermore, the results indicate that any RT dose to the neurogenic niches is correlated with poorer OS in these patients, which is in line with results in our previous study in glioma patients. Based on these data, we suggest that avoidance of these niches is important to extent the OS. However, in a broader NSCLC population with BMs, without any treatment the median survival is only 2 months.^7,29^ When avoidance of the neurogenic niches with radiation is not possible, it is important to carefully consider the potential risks and benefits of RT in patients with BMs from NSCLC. To verify our OS results and explore the effects on neurocognitive functioning in patients with BMs it is necessary to conduct comparable clinical trials prospectively involving larger cohorts of patients suffering from BMs such as the COIMBRA trial.

## Data Availability

The data will be made available upon publication.

## DATA AVAILABILITY STATEMENT

The data will be made available upon publication.

## REFERENCES

1. Nooreldeen R, Bach H. Current and future development in lung cancer diagnosis. Vol. 22, International Journal of Molecular Sciences. 2021.

2. Barnholtz-Sloan JS, Sloan AE, Davis FG, Vigneau FD, Lai P, Sawaya RE. Incidence Proportions of Brain Metastases in Patients Diagnosed (1973 to 2001) in the Metropolitan Detroit Cancer Surveillance System. Journal of Clinical Oncology. 2004 Jul 15;22(14):2865–72.

3. Fenske DC, Price GL, Hess LM, John WJ, Kim ES. Systematic Review of Brain Metastases in Patients With Non–Small-Cell Lung Cancer in the United States, European Union, and Japan. Clin Lung Cancer. 2017 Nov 1;18(6):607–14.

4. Nayak L, Lee EQ, Wen PY. Epidemiology of brain metastases. Curr Oncol Rep. 2012;14(1):48– 54.

5. Sperduto PW, Yang TJ, Beal K, Pan H, Brown PD, Bangdiwala A, et al. Estimating survival in patients with lung cancer and brain metastases an update of the graded prognostic assessment for lung cancer using molecular markers (Lung-molGPA). Vol. 3, JAMA Oncology. 2017. p. 827–31.

6. Mujoomdar A, Austin JHM, Malhotra R, Powell CA, Pearson GDN, Shiau MC, et al. Clinical Predictors of Metastatic Disease to the Brain from Non–Small Cell Lung Carcinoma: Primary Tumor Size, Cell Type, and Lymph Node Metastases. Radiology. 2007 Mar;242(3):882–8.

7. Peters S, Bexelius C, Munk V, Leighl N. The impact of brain metastasis on quality of life, resource utilization and survival in patients with non-small-cell lung cancer. Cancer Treat Rev. 2016 Apr;45:139–62.

8. Hatton N, Samuel R, Riaz M, Johnson C, Cheeseman S, Snee M. A study of non small cell lung cancer (NSCLC) patients with brain metastasis: A single centre experience. Cancer Treat Res Commun [Internet]. 2023 Jan 1 [cited 2023 Jan 11];34:100673. Available from: https://linkinghub.elsevier.com/retrieve/pii/S2468294222001642

9. Goldberg SB, Contessa JN, Omay SB, Chiang V. Lung Cancer Brain Metastases. The Cancer Journal. 2015 Sep;21(5):398–403.

10. Shaw E, Scott C, Souhami L, Dinapoli R, Kline R, Loeffler J, et al. Single dose radiosurgical treatment of recurrent previously irradiated primary brain tumors and brain metastases: final report of RTOG protocol 90-05. International Journal of Radiation Oncology*Biology*Physics. 2000 May;47(2):291–8.

11. McTyre E, Scott J, Chinnaiyan P. Whole brain radiotherapy for brain metastasis. Surg Neurol Int. 2013;4(Suppl 4):S236–44.

12. Brown PD, Ballman K v., Cerhan JH, Anderson SK, Carrero XW, Whitton AC, et al. Postoperative stereotactic radiosurgery compared with whole brain radiotherapy for resected metastatic brain disease (NCCTG N107C/CEC·3): a multicentre, randomised, controlled, phase 3 trial. Vol. 18, The Lancet Oncology. 2017. p. 1049–60.

13. Nagtegaal SHJ, David S, van Grinsven EE, van Zandvoort MJE, Seravalli E, Snijders TJ, et al. Morphological changes after cranial fractionated photon radiotherapy: Localized loss of white matter and grey matter volume with increasing dose. Clin Transl Radiat Oncol. 2021 Nov;31:14–20.

14. Nagtegaal SHJ, David S, Philippens MEP, Snijders TJ, Leemans A, Verhoeff JJC. Dose-dependent volume loss in subcortical deep grey matter structures after cranial radiotherapy. Clin Transl Radiat Oncol [Internet]. 2021 Jan 1 [cited 2023 Mar 28];26:35. Available from:/pmc/articles/PMC7691672/

15. Capilla-Gonzalez V, Bonsu JM, Redmond KJ, Garcia-Verdugo JM, Quiñones-Hinojosa A. Implications of irradiating the subventricular zone stem cell niche. Stem Cell Res. 2016 Mar;16(2):387–96.

16. Achanta P, Capilla-Gonzalez V, Purger D, Reyes J, Sailor K, Song H, et al. Subventricular zone localized irradiation affects the generation of proliferating neural precursor cells and the migration of neuroblasts. Stem Cells. 2012;30(11):2548–60.

17. Wan JF, Zhang SJ, Wang L, Zhao K le. Implications for preserving neural stem cells in whole brain radiotherapy and prophylactic cranial irradiation: A review of 2270 metastases in 488 patients. J Radiat Res. 2013;54(2):285–91.

18. Khacho M, Harris R, Slack RS. Mitochondria as central regulators of neural stem cell fate and cognitive function. Nat Rev Neurosci. 2019 Jan;20(1):34–48.

19. Michaelidesová A, Konířová J, Bartůněk P, Zíková M. Effects of radiation therapy on neural stem cells. Genes (Basel). 2019;10(9).

20. Bruil DE, David S, Nagtegaal SHJ, de Sonnaville SFAM, Verhoeff JJC. Irradiation of the subventricular zone and subgranular zone in high- and low-grade glioma patients: an atlasbased analysis on overall survival. Neurooncol Adv [Internet]. 2022 Jan 1;4(1):1–12. Available from: https://academic.oup.com/noa/article/doi/10.1093/noajnl/vdab193/6506538

21. Oken MM, Creech RH, Davis TE. Toxicology and response criteria of the Eastern Cooperative Oncology Group. pVol. 5, American Journal of Clinical Oncology: Cancer Clinical Trials. 1982. p. 649–55.

22. Radwan AM, Emsell L, Blommaert J, Zhylka A, Kovacs S, Theys T, et al. Virtual brain grafting: Enabling whole brain parcellation in the presence of large lesions. Neuroimage. 2021 Apr 1;229:117731.

23. Park C, Papiez L, Zhang S, Story M, Timmerman RD. Universal Survival Curve and Single Fraction Equivalent Dose: Useful Tools in Understanding Potency of Ablative Radiotherapy. Int J Radiat Oncol Biol Phys. 2008;70(3):847–52.

24. van Gelder BM, Tijhuis MAR, Kalmijn S, Giampaoli S, Kromhout D. Decline in Cognitive Functioning Is Associated with a Higher Mortality Risk. Neuroepidemiology [Internet]. 2007 [cited 2023 Mar 28];28(2):93–100. Available from: https://www.karger.com/Article/FullText/98552

25. Gondi V, Pugh SL, Tome WA, Caine C, Corn B, Kanner A, et al. Preservation of Memory With Conformal Avoidance of the Hippocampal Neural Stem-Cell Compartment During Whole-Brain Radiotherapy for Brain Metastases (RTOG 0933): A Phase II Multi-Institutional Trial. Journal of Clinical Oncology. 2014 Dec 1;32(34):3810–6.

26. Valiyaveettil D G A, Malik M, Eaga P, Ahmed SF, Joseph D. “A prospective study of assessment of neurocognitive function in illiterate patients with gliomas treated with chemoradiation”: Assessment of neurocognitive function in gliomas. Cancer Treat Res Commun. 2021 Jan 1;26:100288.

27. Hoshide R, Jandial R. The role of the neural niche in brain metastasis. Clin Exp Metastasis. 2017;34(6–7):369–76.

28. Oskan F, Ganswindt U, Schwarz SB, Manapov F, Belka C, Niyazi M. Hippocampus sparing in whole-brain radiotherapy: A review. Strahlentherapie und Onkologie. 2014;190(4):337–41.

29. Jeene PM, de Vries KC, van Nes JGH, Kwakman JJM, Wester G, Rozema T, et al. Survival after whole brain radiotherapy for brain metastases from lung cancer and breast cancer is poor in 6325 Dutch patients treated between 2000 and 2014. Acta Oncol (Madr). 2018 May;57(5):637–43.

